# Serum mtDNA DAMP abundance, fragmentation and heteroplasmic variants associate with Acute Respiratory Failure outcome: A secondary analysis of study NCT00976833

**DOI:** 10.1101/2025.08.26.25334376

**Authors:** Grant T. Daly, Emily M. Hartsell, Viktor M. Pastukh, Justin T. Roberts, Adeyeye I. Haastrup, Lina D. Purcell, Madhuri S. Mulekar, D. Clark Files, Peter E. Morris, Mark N. Gillespie, Raymond J. Langley

## Abstract

**Background:** Serum mitochondrial DNA (mtDNA) fragments act as proinflammatory damage-associated molecular patterns (DAMPs), and have been linked to outcomes in critical illness. However, their prognostic value remains uncertain, possibly due to confounding nuclear mitochondrial insertions (NUMTs) which obscure both quantitation and variant detection.

**Methods:** Using a targeted deep sequencing and bioinformatics workflow, we created filtering strategies to minimize NUMT-related artifacts. To evaluate the method, we performed a secondary analysis of serum samples collected from NCT00976833, a study of acute respiratory failure patients. By modeling DNA insert size distributions, we excluded likely NUMT-derived DNA fragments based on their size, improving the accuracy of mtDNA DAMP fragmentomic analysis. To improve variant detection, we introduced a novel “read mismatch percentage” metric to identify NUMT-induced chimeric read pairs, enabling identification of mtDNA variants.

**Results:** Mean NUMT-depleted, but not raw, mtDNA insert size was lower in non-survivors. Short DNA inserts (<150 bp) displayed little NUMT contamination, and their abundance and size correlated with mortality more strongly than total mtDNA abundance. Sequence variants were called and some associated with survival and post-acute quality of life. Variant m.1,719G*>*A, found in small humanin-like 3 (*MT-SHLP3*), associated with survival. Other variants associated with overall poor outcome (non-survival or poor QoL). Two noncoding variants previously associated with low VO2 max and coronary artery disease (m.295C*>*T and m.462C*>*T) also associated with poor outcome in the present study. Two *MT-ND5* variants m.13,708G*>*A (a missense variant previously implicated in kidney dysfunction) and m.12,612A*>*G (a synonymous variant previously associated with coronary artery disease) also associated with poor overall outcome.

**Conclusions:** Our results addressed limitations of standard qPCR-based methods for the study of mtDNA DAMPs. Beyond addressing confounding NUMT, the method identified fragmentomic and variant associations overlooked by qPCR. Cell-free DNA fragmentomic and variant information are well-established biomarkers for cancer, and this method could facilitate similar patient-specific biomarkers in the context of critical illness. The method is composed of commercially available reagents and open source software, which could additionally promote adoption and reproducibility.

## Introduction

Mitochondrial damage is centrally implicated in the pathogenesis of critical illness and the consequences of severe conditions in human patients, a conclusion supported by findings from relevant animal models [1–5]. Such mitochondrial injury is likely driven by elevated oxidative stress, a phenomenon extensively documented across various disease states [6–8]. Beyond their traditional bioenergetic roles, mitochondria also serve as platforms for assembling proinflammatory signalosomes and releasing inflammatory mediators [9]. This dual functionality provides a plausible mechanism by which mitochondrial damage may orchestrate the cascade of events leading to critical illness or injury.

Over a decade ago, studies by Zhang et al. established that traumatic injury leads to the release of mitochondrial DNA (mtDNA) fragments, now classified as mitochondrial DNA damage-associated molecular patterns (mtDNA DAMPs), into the systemic circulation, where they can trigger a Toll-like receptor 9 (TLR9)-dependent immune response [10, 11]. Notably, mtDNA DAMPs themselves can induce further oxidative damage to mtDNA, potentially initiating a self-sustaining inflammatory feedback loop [12]. Repeated findings of mitochondrial damage and associated inflammatory complications in critically ill and injured patients have motivated extensive research into the relationship between circulating mtDNA DAMP levels and clinical outcomes. A systematic review found that most studies report positive associations between mtDNA abundance and mortality, but that the prognostic utility of mtDNA DAMPs is still uncertain [13].

The limited prognostic capability of circulating mtDNA assays may stem in part from interference by nuclear mitochondrial DNA insertions (NUMTs) [13]. Quantitative real-time PCR (qPCR) is the standard approach to quantify mtDNA DAMPS. However, qPCR suffers from several methodological constraints. Firstly, no standardized protocols exist, particularly concerning primer design [13] Proper primer design is crucial to avoid amplification of NUMT contaminants, which can obscure accurate quantification and result in misclassification of mtDNA sequence variants within circulating DAMPs [14, 15]. Secondly, qPCR targets only predefined mtDNA amplicons, thereby neglecting fragmentomic characteristics, such as those in cell-free prenatal and cancer DNA studies [16]. Structural characteristics are especially relevant for mtDNA DAMPs, as DNA fragment length and CpG structural context are known to impact their affinity for TLR9 [13, 17]. Furthermore, qPCR cannot detect broad spectrum mtDNA sequence variants, a significant limitation given the multitude of pathologies associated with mtDNA mutations [18]. Mitochondrial DNA mutations are well-known causes of pathology, and are increasingly appreciated for their diagnostic utility in cancer [19]. It stands to reason that mtDNA variants could similarly associate with critical illness outcome.

To address these NUMT-related methodological shortcomings, we refined our previously described deep sequencing approach for mtDNA to judiciously account for annotated for annotated and non-annotated polymorphic NUMTs thereby enabling more precise quantification of mtDNA DAMP abundance and fragmentomic features as well as facilitating detection of low-frequency mtDNA variants [18]. To assess the potential value of this improved approach, we analyze samples from the TARGET study (NCT00976833), a randomized clinical trial evaluating early physical therapy in patients with acute respiratory failure (ARF) [20–22]. We assessed mtDNA DAMP abundance, DNA insert size, and heteroplasmic variants to ascertain whether these molecular features could differentiate survivors from non-survivors and further distinguish survivors with favorable outcomes from survivors with unfavorable physical function trajectories.

## Results

### Patient Demographics & Study Inclusion Criteria

This study utilized patient samples enrolled in the Standardized Rehabilitation for Intensive Care Unit (ICU) Patients With Acute Respiratory Failure trial (NCT00976833), a single center, single blind, randomized control study designed to determine the effects of physical therapy initiated in ARF [20–22]. The present study sought to contrast attributes of serum mtDNA in ARF survivors (*n* = 36) versus non-survivors (*n* = 36), all samples taken upon ICU admission (Table S1a), and in survivors with good ( *n* = 23) versus poor (*n* = 26) quality of life (QoL) (Table S1b). For both the survival and QoL arms of our studies, serum was isolated from venous blood samples obtained upon ICU admission. In the QoL study, admission and discharge samples were available for 28*/*49 patients, whereas only one timepoint was available for the remaining patients (Table S1). QoL was measured in hospital using the Short Physical Performance Battery (SPPB) trajectory model introduced by Gandotra and colleagues [21], which demonstrated that SPPB assessed in hospital are predictive of long-term SPPB scores up to six months following the hospital stay. For the survival study, the average length of hospital stay was 14 *±* 19 (sample mean *±* stdev.) days and 13 *±* 10 days until death for non-survivors (Table 1). For the QoL study, average length of stay was 16 *±* 16 days (Table 1).

**Table 1.**
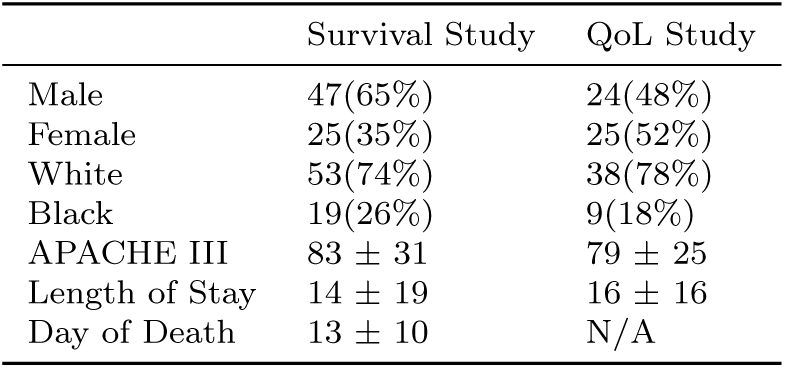
Demographics and clinical parameters for survival study and quality of life (QoL) study.

|  | Survival Study | QoL Study |
| --- | --- | --- |
| Male | 47(65%) | 24(48%) |
| Female | 25(35%) | 25(52%) |
| White | 53(74%) | 38(78%) |
| Black | 19(26%) | 9(18%) |
| APACHE III | $83 \pm 31$ | $79 \pm 25$ |
| Length of Stay | $14 \pm 19$ | $16 \pm 16$ |
| Day of Death | $13 \pm 10$ | N/A |

### Approach to identify polymorphic NUMTs in the mtDNA read pool

A major challenge associated with deep sequencing analysis of the serum mtDNA pool is identifying and mitigating contaminating NUMTs from the data analysis. Not only do these nuclear DNA (nDNA) fragments have the potential to introduce errors in quantification of mtDNA DAMPs and assessment of fragment lengths, but they also obfuscate identification of authentic mtDNA variants due to sequence homology between mtDNA and NUMTs. More problematic are polymorphic NUMTs, as they are common in the population and absent from the reference genome [14].

We developed a bioinformatic protocol to identify NUMTs and minimize the impact of NUMT on the quantification of mtDNA DAMP abundance, DNA insert size, and variant detection. Our approach was based on the premise that sequenced NUMT reads, originating from the nuclear genome, should appear as pseudo-mtDNA fragments approximately the length of nucleosome-associated DNA (*≈* 166 *−* 167 bp) [23–25]. Illumina library preparation blunts any 3’ or 5’ overhangs from the DNA fragment, so we measure “insert size” in place of fragment size (see Methods for details). Consistent with our hypothesis, insert size distribution analysis of patient mtDNA sequences ( Figure 1a and Figure S1) revealed a prominent peak conspicuously similar to the length of DNA wrapped around the size of mononucleosomes plus a *≈* 20 bp linker sequence [24, 26]. In contrast, the insert size curve for mtDNA DAMPs was broad and right-skewed.(Figure 1a; Figure S1). Mitochondrial DNA insert size distribution curves for 14 % of samples (21 out of 149) displayed steep peaks similar to those of annotated NUMTs in both size and shape (Figure S1), suggesting that some polymorphic NUMTs had escaped bioinformatic filtering. This was likely due to their absence from the reference genome and high sequence homology with authentic mitochondrial DNA. Supporting this, more than 99 % of humans carry at least one polymorphic NUMT, and approximately 1 in 8 individuals harbor an ultra-rare NUMT, defined as being present <0.1 % of the population [14].

**Fig. 1.**
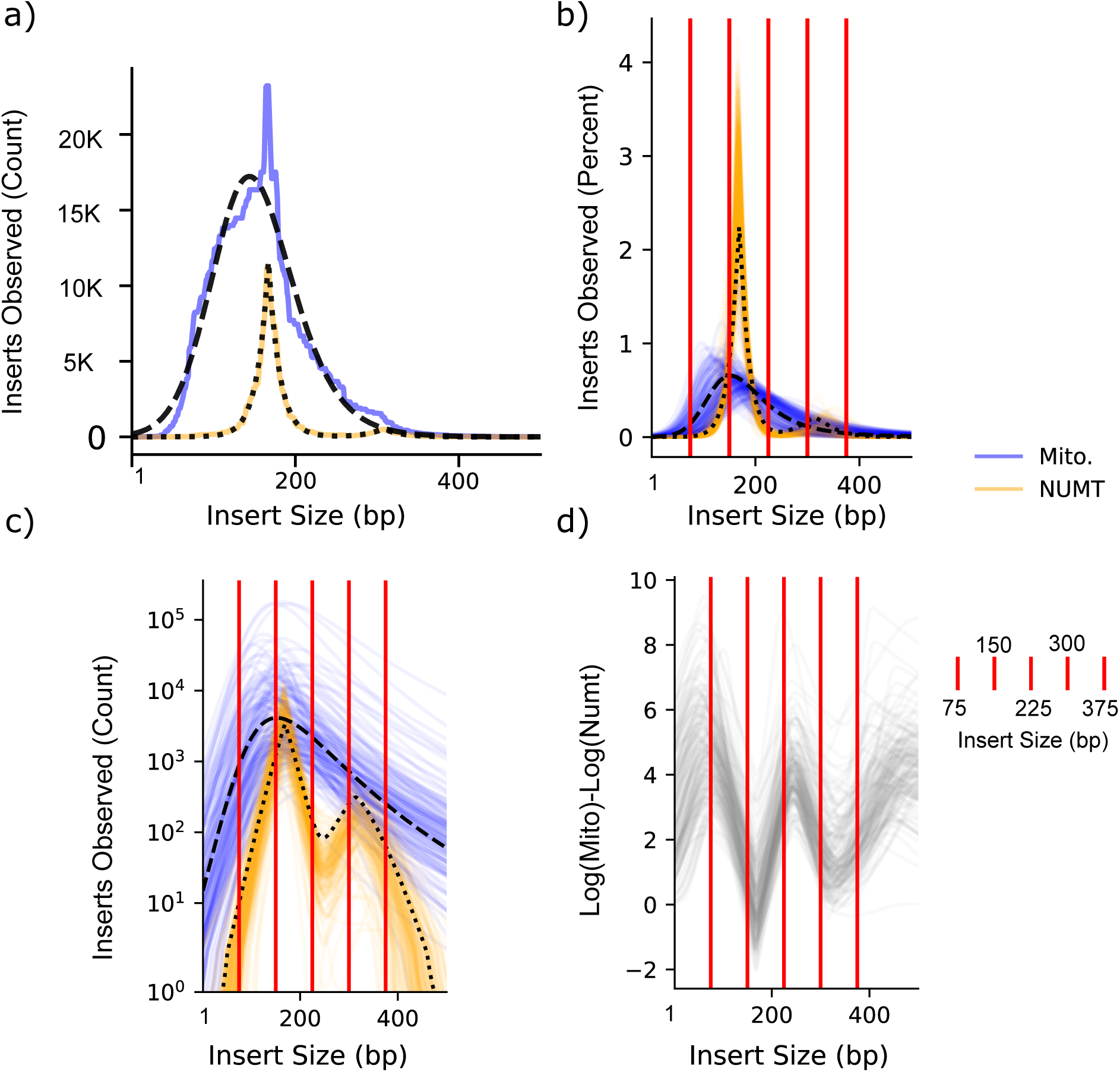
Insert size distributions. (A) Insert size distributions scaled as number of inserts and percentages for one representative sample with overlayed fitted curves for mitochondria (raw data orange, dashed black fitted curve) and NUMT (raw data yellow, dotted black fitted curve). (B and C) Fitted mtDNA and NUMT curves for all samples displayed as percent (B) and counts (C), plotted on logarithmic scale with median fitted curve by Monte Carlo sampling overlayed as dashed black line (mitochondria) and dotted black line (NUMT). (D) The log difference between mitochondrial and NUMT reads counts observed by insert size. Red vertical lines demarcate 75 bp bins used to create insert abundance metric.

### Optimizing characterization and quantitation of mtDNA DAMPs

Based on the realization that annotated NUMTs are prominent in specific size ranges, we determined if mitochondria-aligned reads in those specific size ranges were relatively enriched in polymorphic NUMTs. This information, by exclusion, highlights those ranges that were relatively NUMT free; providing a guide for accurate quantitation of mtDNA DAMPs in patient populations. First, to delineate insert sizes ranges containing high levels of putative NUMTs we modeled insert size distributions for individual patients’ total mtDNA and for annotated NUMTs fitting to a Skew Student’s t distribution and a Generalized Normal distribution, respectively (see Methods for details). Next, to estimate insert size parameters, we pooled sample results among all patients and performed a simple Monte Carlo simulation (10,000 samples with replacement; see Methods for details), measuring the 10th, 50th, and 90th quantile of inserts observed within each draw [27]. The median value of all 10,000 draws was chosen as a point statistic, and is presented in Table S3 for mitochondria, mononucleosomes, and dinucleosomes. Notably, the 50th percentile values for mononucleosome and dinucleosome peaks (median 167 bp and 319 bp, respectively) closely matched reported values for nucleosome-bound nDNA [24]. The key finding is that the vast majority (80 %) of mononucleosomal NUMT sequences are between 145-189 bp (Table S3), consistent with prior observations that cell-free nDNA fragments are rarely observed below 100 bp [25]. If mitochondrial copy number was sufficiently high compared to nDNA copy number, it would imply that polymorphic NUMTs are not a major concern for quantitative mtDNA analyses. Figure 1c displays insert size distributions as raw read counts, displayed on a logarithmic scale. The peak mononucleosomal NUMT’s counts did intersect with many of the mitochondrial fitted distributions (Figure 1c). To verify that individual samples had insert size ranges with mitochondrial and NUMT abundance, the difference between mitochondrial and NUMT counts for each sample is displayed as Figure 1d. As expected, the lowest differences are observed at putative mononculeosome and dinucleosome peaks. Taken together, these findings suggest that polymorphic NUMT are most likely to present in well-defined insert size ranges corresponding to the sizes of mononucleosomes and dinucleosomes, and that mtDNA inserts shorter than 150 bp are essentially devoid of NUMTs and enriched in mtDNA (Figure 1b-d, Table S3). We next determined how NUMT contamination in various size ranges of total DNA inserts would impact determination of authentic mtDNA DAMPs abundance. Our previous work calculated abundance as normalized “coverage” (mitochondrial genome coverage*/*NUMT coverage) [18]. However, ligands that activate PRRs are known to have affinity for short fragments, which is not explicitly measured by coverage or typical qPCR methods [13, 18, 28]. Therefore, we also calculated abundance based on the absolute number of inserts (i.e., fragments) of a given size range normalized to NUMTs.

Because NUMTs are present at very low levels at insert sizes less than 150 bp, we postulated that quantitation mtDNA DAMPs at insert sizes in that size range will provide the most accurate determination of abundance (Figure 1b-d). If true, then abundance from inserts in this size range would be expected to correlate poorly with abundance determined using insert sizes that contain significant NUMT contamination (i.e, inserts *>*150 bp). To test this idea, we performed correlation analyses between normalized coverage, abundance for all insert sizes (included insert size 75-374 bp), and abundance binned by insert size (75-149 bp, 150-224 bp, 225-299 bp, 300-374 bp); see vertical red lines in Figure 1b-d, normalized to mean NUMT inserts from 75-374 bp Consistent with our expectation, we found that all inserts were highly correlated with normalized coverage (*ρ* = 0.96, Pearson), indicating that both metrics similarly estimate combined mtDNA DAMP and polymorphic NUMT abundance (Figure S2). Also as expected, inserts less than 150 bp are moderately correlated with normalized coverage (*ρ* = 0.75) and all insert abundances (*ρ* = 0.89; Figure S2). Note that fragments less than 74 bp are largely undetectable by the sequencing method used herein. Finally, longer insert size bins (225-299 bp and 300-374 bp) were poorly correlated with short inserts (*ρ* = 0.56 and 0.23, respectively), further implying that NUMT contamination may reduce the accuracy of plasma mtDNA DAMP abundance, especially for the longer size ranges. These findings demonstrate the ability to identify a range of insert sizes in which NUMT contamination is negligible, thereby minimizing the confounding effects of polymorphic NUMTs and enabling more accurate quantification of mtDNA DAMPs. Below, we apply this workflow to evaluate its utility in the early discrimination between survivors and non-survivors of ARF, and among survivors, to distinguish between those with good versus poor QoL (e.g., physical function).

### mtDNA DAMP parameters as predictors of acute and post-acute outcomes

We applied Kruski’s “BEST” (Bayesian Estimation Supersedes the t Test) modeling framework on log-transformed coverage or abundance data with a Normal distribution likelihood function (see Methods for details) [29]. Figure 2 compares coverage and abundance for the various insert size bins between survivors and non-survivors. Figure 2a shows scattergrams and the “BEST” 95 % highest density interval (HDI) of the difference between the means. As expected, the shortest inserts were most associated with survival, and the difference was slightly lower for the 150-225 bp bin predicted to have high NUMT but also high mitochondrial abundance. No difference was detected for the 225-299 and 300-374 bp bins, which were predicted to have appreciable dinucleosomal NUMT abundance and comparatively low mitochondrial abundance.

**Fig. 2.**
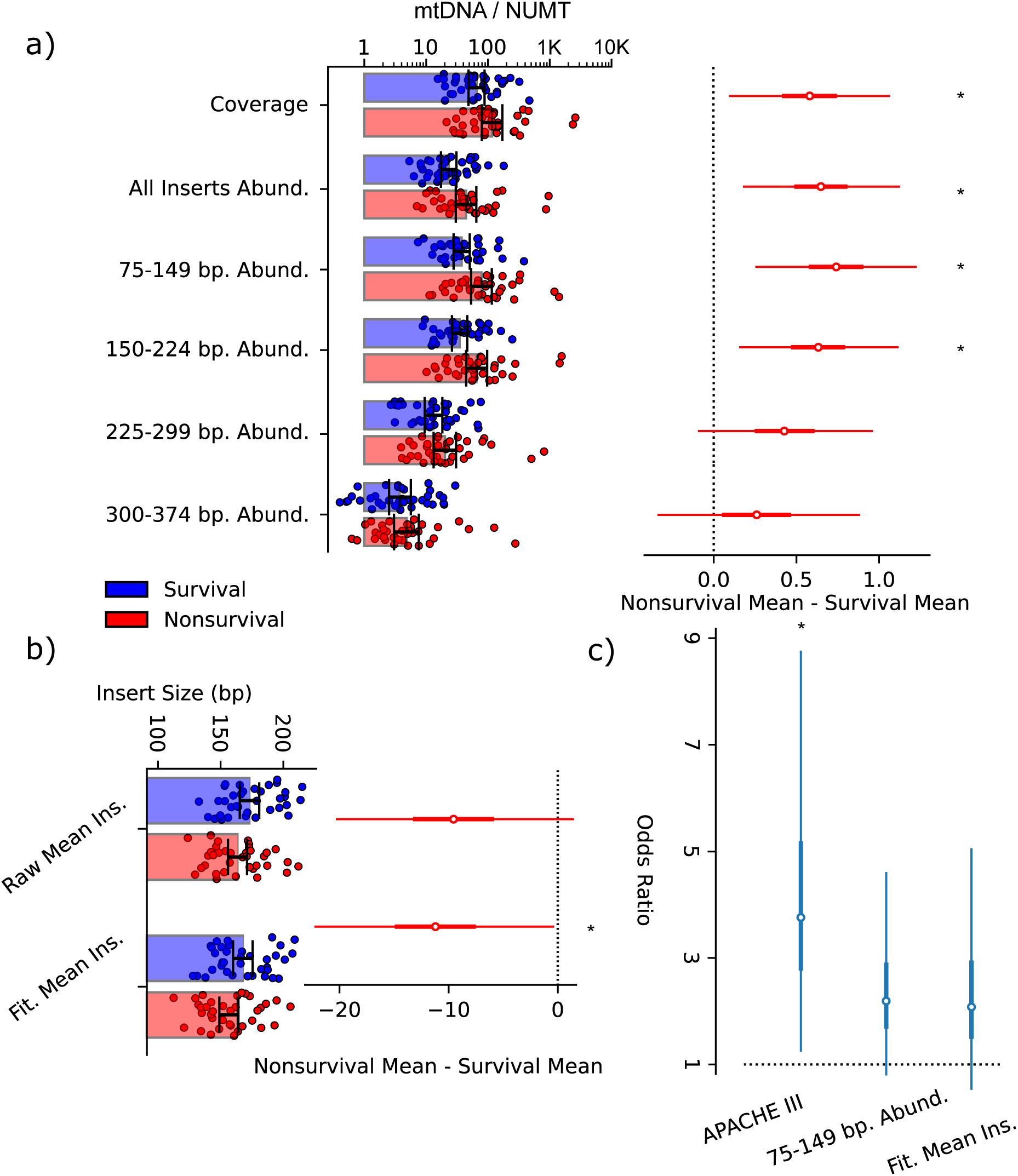
mtDNA features and acute outcome (A) Posterior distribution of coverage and abundance plotted as individual points with bars displaying 95 % HDI and mean differences by acute outcome, displaying 95 % HDI as horizontal bar and difference “0” as dotted vertical line. (B) Mean raw or fitted fitted mtDNA insert size mean for individual data points, with 95 % HDI bars. 95 % HDI for the difference in mtDNA mean insert size by acute outcome (nonsurvival - survival), displaying 95 % HDI as horizontal red bars and difference “0” as dotted vertical line. (C) Odds ratio for logistic regression model predicting non-survival, comparing the 75th to 25th percentile for the denoted parameter. Horizontal bars display 95 % HDI’s of the denoted odds ratio, and the vertical bar placed at odds ratio “1” signifies no difference from random chance.

Because the data approximated a Gaussian distribution, we applied a Student’s t likelihood function to derive a corresponding “BEST” model for insert sizes (Figure 2b). Mean insert size was lower in non-survivors compared to survivors for the fitted (but not raw) data (Figure 2b). This was consistent with our previous study of moderately injured patients where shorter insert lengths associated with post-traumatic complications [18].

We next determined if our new methods for quantifying mtDNA DAMP features improved prediction of acute patient outcomes relative to the established APACHE III score.(Knaus et al., 1991) A Bayesian logistic regression model was created with the previously mentioned top performing measures 75-149 bp, fitted mean insert size, and APACHE III. Odds ratios by quartile are displayed in Figure 2c. As expected, APACHE III 95 % HDI was higher than 1, although it displayed a broad distribution. Abundance and mean insert size did not exceed 1 but displayed considerably less variance. This is consistent with previous studies that mtDNA DAMPs features are moderately predictive of patient outcomes [13].

Post-intensive care syndrome (PICS) complicates long-term outcomes in survivors of critical illness, such as ARDS, pneumonia, and sepsis [30]. Two prior studies using qPCR methods presented discordant results as related to mtDNA DAMPs and the association between PICS, with one showing no association [31]; whereas the second reported a moderate association [32]. Here, focusing on ARF survivors, we sought to determine if properties of mtDNA DAMPs associated with physical function trajectories. Physical function was measured using the Short Physical Performance Battery (SPPB), as previously described [21, 22]. The SPPB test has emerged as a useful surrogate for physical function [33, 34]. The test, as applied herein, includes an objective assessment of physical function as determined by gait speed, balance, and lower extremity strength [34]. Scores range from 0-12, with 0-3 describing significant physical function disability, 4-6 low physical function, 7-9 intermediate physical function, and 10-12 high physical function [34]. In the current study, we defined patients with “low” QoL as having an SPPB score <6, while values *>*7 identified patients with good QoL.

A “BEST” Bayesian model, similar to that described above, was extended to include time (admission and discharge) as an additional factor. To account for repeated measures, admission and discharge were given a correlation term and missing values were imputed from the Markov Chain Monte Carlo (MCMC) sampling (See Methods for details). Abundance scattergrams are provided for coverage and 75-149 bp abundance in Figure 3a, and corresponding scattergrams for raw and fitted mean insert size are presented in Figure 3b. Mean differences were called between discharge and admission (for both good and poor QoL; Figures S3b and S4b) and between poor and good QoL (for both discharge and admission; Figures S3c and S4c) for both 75-149 bp abundance and coverage (Figure S3) and raw and fitted insert size (Figure S4). No differences in means by time or QoL met the 95 % HDI cutoff for abundance or insert size.

**Fig. 3.**
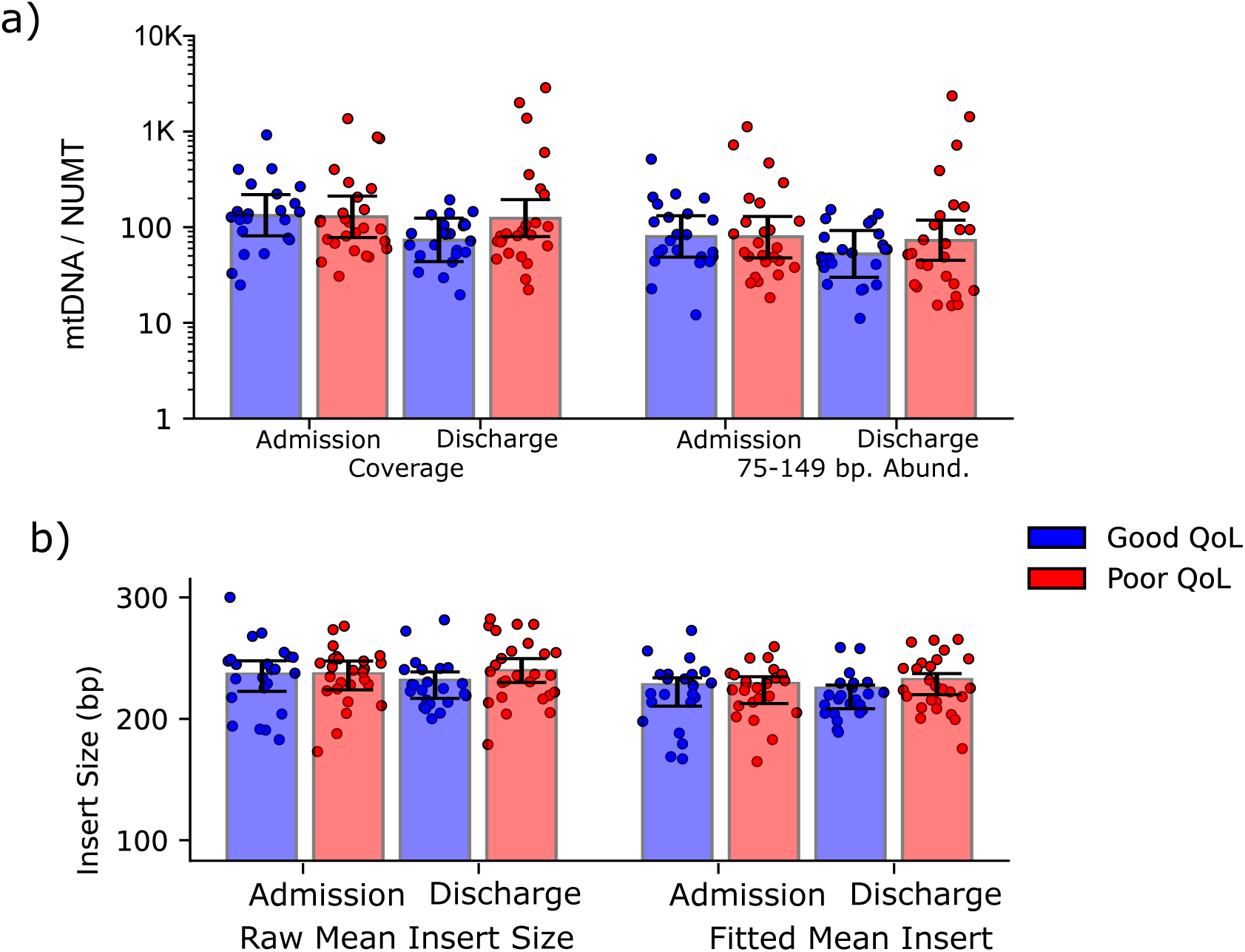
mtDNA parameters by QoL and time (A) Data points and 95 % HDI for coverage and 75-149 bp inserts. (A and B) 95 % HDI of differences in coverage or abundance by (B) time (discharge - admission) or (C) QoL (poor - good). Difference “0” is displayed as a dotted vertical line.(B) Points and 95 % HDI for raw and fitted mean insert size. (A and B) 95 % HDI of insert size differences by time (B) (discharge - admission), and QoL (C) (poor QoL - good QoL). Difference “0” displayed as dotted vertical line.

### Heteroplasmic variants associate with acute and overall post-acute outcomes in ARF

Variant signatures in DNA obtained from liquid biopsies can serve as diagnostic markers for various diseases, particularly cancers [16]. Although mtDNA variants have been associated with several disorders, detecting these variants from liquid biopsies remains challenging due to their low abundance relative to nDNA and, more critically, contamination by NUMTs. A particularly difficult issue arises from polymorphic NUMTs, which can persist despite bioinformatic filtering, further complicating accurate mtDNA analysis. Somatic NUMTogenesis has also been detected, which suggests that polymorphic NUMT’s are a patient-specific feature masquerading as authentic mtDNA variants [15, 35].

In light of this, we devised a method to facilitate polymorphic NUMT detection and thereby account for associated pseudo-variants. The “Dinumt” method previously identified NUMT based on chimeric read pairs which aligned to both the nuclear and mitochondrial genomes [15, 36]. We adapted this strategy to develop a “read mismatch percentage” defined as the number of variant bases from chimeric read pairs divided by the total number of variant bases from all reads: read mismatch % = (chimeric variant*/*chimeric variant + mtDNA-aligned variant). This was designed to directly measure the percentage of chimerism for any given alternate allele, and thus directly measure the impact of NUMT on variant calling. We present mismatch percentages in terms of their position within the mitochondrial genome in Figure S5a. There are regions across the genome that are conspicuous for clusters of read mismatch percentages. We next inspected representative regions with high mismatch percentages using IGV [37] (Figure S5b), which clearly demonstrated the insertion sites in both the nuclear and mitochondrial genomes, and which allowed for the identification of potential mtDNA pseudo-variants.

Traditionally, heteroplasmic variants have been defined by the number of reads harboring a variant relative to the total number of reads, termed the variant allele frequency (VAF). Accordingly, we wondered how variants defined by their VAF related to the mismatch percentages described above. As shown in Figure 4 this relationship was surprising: some variants characterized by high VAFs also displayed high mismatch percentages. In contrast, some low VAF variants had extremely low mismatch percentages, suggesting that they were authentic variants despite the low VAF. These findings call into question the validity of using a sole VAF threshold to call heteroplasmic variants.

**Fig. 4.**
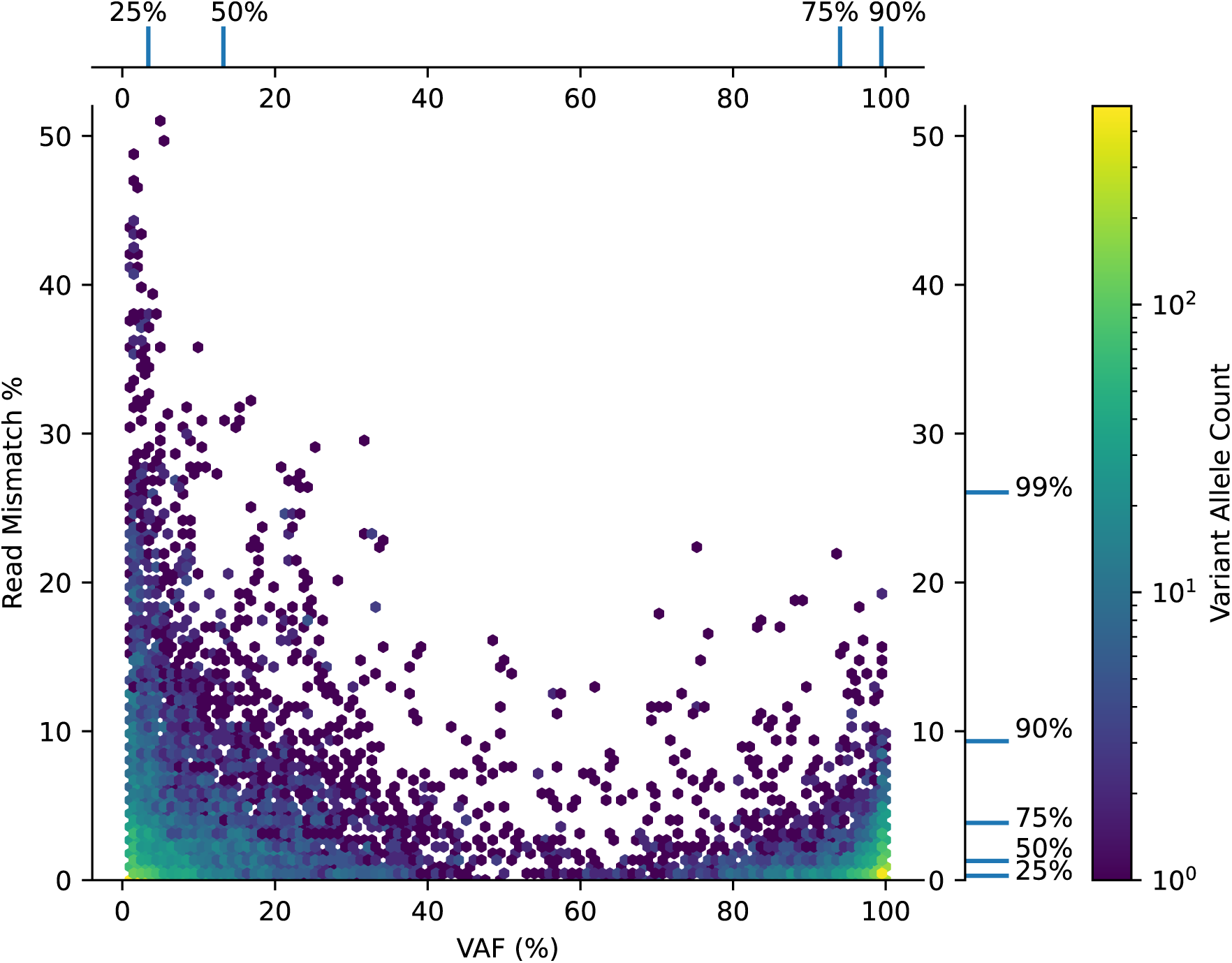
Read mismatch percent. Read mismatch % (one read in a read pair mapped to the mitochondrial genome with the mate read mapped to the nuclear genome) vs. variant allele fraction (VAF). Selected quantiles are plotted in the margin, and the color scale corresponds to the number of loci observed.

To remove likely NUMT with high mismatch percentages while not removing likely authentic high-VAF variants which happened to align chimerically to a reference NUMT, our assessment of mtDNA variants in ARF incorporated both the mismatch percentages and the variant allele frequency (VAF), with the former used as the initial screen for authentic variants. A variant was considered authentic and eligible for further analysis if its mismatch percentage was below the 90th percentile of all observed values–approximately a 10 % mismatch (Table S8). For variants meeting this criterion, we applied VAF thresholds to compare outcome groups. Traditionally, a fixed variant allele frequency (VAF) cutoff (e.g., 25 %) is used to define high-quality variants. However, this can be misleading when comparing groups, as small, biologically insignificant differences (e.g., 24 % vs. 26 % VAF) may disproportionately affect how variants are distributed between outcome groups. To address this, we aimed to identify variants showing large VAF differences between clinically relevant outcomes. When searching for variants that associate with a positive outcome (e.g., survivors), we required VAF *>* 25% in surviving patients and < 1% in non-survivors. The converse relationship was used when searching for variants associated with negative outcomes (e.g., non-survivors).

This analytical approach was applied to two specific comparisons, 1) surviving versus non-surviving patients, and 2) all negative outcomes (i.e. death or survival with poor physical function) versus good outcomes (survival with good physical function). All patients were pooled for these analyses because variants called at VAF 25 % were likely germline, so the strategy was likely to be robust to batch effects. The risk of batch effects in the continuous-valued abundance and insert size analyses had been our motivation to separate the for survival and QoL analyses. Association p-values and ratios for these comparisons are displayed in Figure 5. Variants with *p <* 0.05 and minimum ratio 5 are annotated in Figure 5 and more information about them is provided in Table S9, with added annotations from the ENSEMBL Variant Effect Predictor API [38], Mitomap [39] database, and IGV [37].

**Fig. 5.**
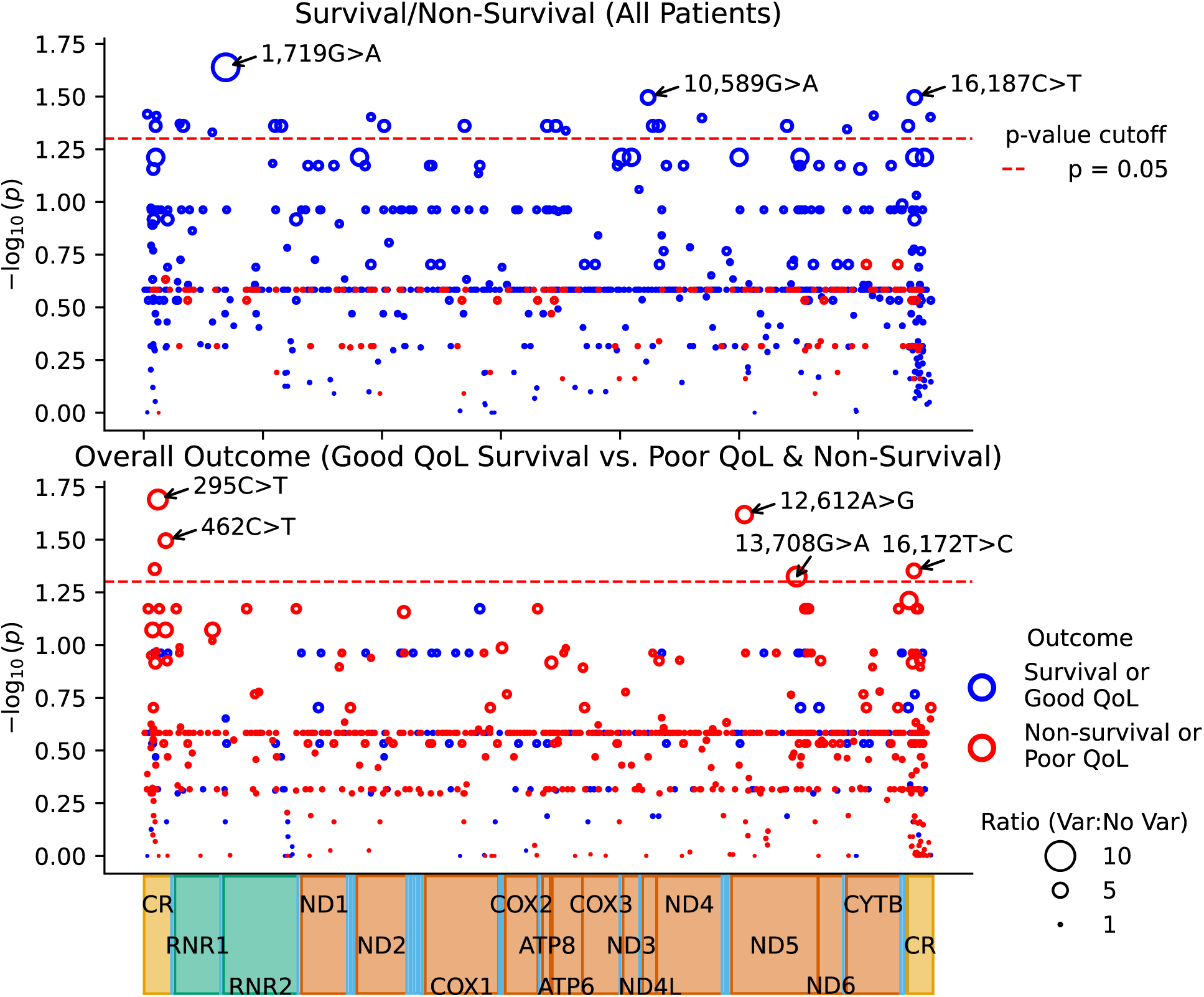
Outcome associations by Permutation test. Permutation test results for the number of patients with a variants at minimum 25 % VAF threshold for the outcome compared to the number less than 1 % VAF in the outcome being compared against. Data are displayed as*−* log 10(*p*) by mitochondrial coordinate. Data are sized by the variant ratio. Variants *p <* 0.05 and with minimum ratio 5 are annotated with position and variant type.

For survival (*n* = 85) vs. non-survival (*n* = 36), there were three variants significantly associated with survival. Perhaps most interesting, m.1,719G*>*A is predicted to be a synonymous variant in small humanin-like 3 (*MT-SHLP3*; *p* = 0.033, and a 10:1 ratio of survival to non-survival (Table S9a). In terms of overall poor outcome, (non-survival or survival with poor QoL, *n* = 78) and overall good outcome (survival with good QoL; *n* = 43), five variants were significantly associated with the overall poor outcome at *p <* 0.05 (Table S9b). Two control region variants (m.295C*>*T and m.462C*>*T) have previously been associated with low VO2 max [40, 41] and coronary artery disease [42]. We also observed two protein coding variants which associated with overall poor outcome in the *MT-ND5* gene, a synonymous variant m.12,612A*>*G) previously associated with coronary artery disease [42] (*p* = 0.024), and m.13,708G*>*A, a missense variant at *p* = 0.047 which was previously associated with kidney dysfunction [43] (*p* = 0.047).

## Discussion

Accumulating evidence establishes that mitochondrial dysfunction is a contributor to poor outcomes in ARDS, sepsis, and pneumonia [4, 5, 28, 44–46]. More recent studies have shown that mtDNA DAMPs in cell-free plasma or serum are associated with poor outcomes, pointing to the prospect they are a surrogate biomarker of mitochondrial dysfunction in critical illness and injury. This is supported by involvement of mitochondria in the activation of the innate immune response. However, despite these provocative observations, AUROC analyses generally indicate that mtDNA DAMP abundance is not a particularly robust predictor of outcomes [13]. These findings were recapitulated in the present study. Several factors may explain the limited utility of mtDNA DAMPs as a single-molecule biomarker. Chief among these limitations could be the presence of contaminating NUMTs, which interfere with accurate determinations of mtDNA DAMP abundance analysis [13, 47]. Contaminating NUMTS may also confound the utility of other mtDNA DAMP parameters, such as insert size, which because it requires Illumina sequencing has been infrequently reported [18, 25]. The use of plasma heteroplasmic mtDNA variants as predictors of outcomes and severity also has rarely appeared in the literature despite the emerging use of nDNA as a liquid biopsy substrate [18, 25, 48]. Here, use of serum mtDNA DAMPs for heteroplasmic variant analysis is particularly limited by the confounding impact of NUMTs, as NUMT could present at similar VAF as potential tissue-specific heteroplasmic variants dynamically mobilized during critical illness. Thus, a central goal of the current work was to develop a strategy to reduce the impact of NUMTs in the quantification and structural analysis of mtDNA DAMPs.

We found that mtDNA and nDNA have unique fragmentation patterns that offer clues to differentiate NUMTs from authentic mtDNA. NUMTs tend to appear as fragments clustered around two distinct peaks, the first, narrow peak at 1̃67 bp is consistent with a single wrap of DNA around a mononucleosome (Figure 1), and another broader peak at 3̃30 bp, likely composed of dinucleosomes. In contrast, authentic mtDNA, because it is not protected by histones, exhibits a much broader insert size distribution. Despite bioinformatic filtering of annotated NUMTs, approximately 14 % of the patient samples exhibited small but distinct peaks centered around 167 bp in the insert size distribution graphs, pointing to the prospect that these peaks represented polymorphic NUMT sequences contaminating the mtDNA-aligned read pool.

We used this improved strategy to delineate NUMTs and increase the accuracy of mtDNA DAMP quantification. Based on the above contention that the small size range is almost entirely NUMT-free, we used this insert size range as the “gold standard” for mtDNA quantitation. We also demonstrated that the raw number of NUMT reads was similar to the levels of mtDNA for many samples, particularly at the mononucleosomal and dinucleosomal insert size range. We next performed a correlation analysis between this insert range and bins of longer insert size ranges, The 75-149 bp insert size abundances only modestly correlated with coverage, and correlated poorly with abundances of longer insert sizes, where mitochondrial DNA abundance tailed off and was comparable to dinucleosomal NUMT abundance. We further validated the prospect that insert size dependent differences in abundance reduced the utility of mtDNA DAMPs as a biomarker. As predicted, the largest abundance difference between non-survivors and survivors was observed in the 75-149 bp bin. The difference was slightly lower for the 150-224 bp bin, where NUMT levels were predicted to be the highest, but which also contained high authentic mitochondrial DNA abundance. No difference existed by survival status for the 225-299 and 300-374 bp bins, which was at the range where mitochondrial DNA abundance tailed off and became comparable to dinucleosomal NUMT abundance. Overall, a low mtDNA/NUMT ratio, along with significant polymorphic NUMT contamination, may impact the accuracy of abundance quantification, suggesting that the NUMT-depleted insert size range may be required for the most reliable measurement.

A final goal was to optimize identification of mtDNA variants in cell-free plasma or serum and to determine their association with outcomes of ARF. In this context, only a handful of papers have focused on the mitochondrial genome as a substrate for liquid biopsy [48, 49]. These reports typically utilized a single VAF threshold to call variants. However, a single VAF threshold can be misleading, since small, insignificant differences may disproportionately influence the distribution of variants across outcome groups. In addition, our data demonstrates that we can identify polymorphic NUMT pseudo-variants by their read mismatch percentage. Occasionally, likely NUMT were even observed at VAFs greater than 25 %. Accordingly, we propose a two-step strategy. The first applies the mismatch percentage to filter out suspicious variants prior to subsequent comparisons. Second, we used two VAF thresholds in mtDNA variant selection to allow for more nuanced, clinically relevant analyses. Specifically, variants associating with a positive outcome (e.g., survivors) were required to display a 25 % minimum VAF, while the same variant associated with a negative outcome (e.g., death) is required to be present at less than 1 % VAF. The converse relationship was used when searching for variants associated with negative outcomes.

We applied these methods to quantify mtDNA DAMPs and variants in patients with ARF, specifically survivorship and QoL. In terms of the former, like most other reports, our analysis revealed that non-survivors displayed a higher abundance of cellfree mtDNA DAMPs in serum, while fitted insert size mean was lower in non-survivors, consistent with our previous patients with moderately injured trauma patients [18]. Relative to QoL, as defined by the SPPB physical function scores, no differences met our thresholds for abundance or insert size. We next interrogated mtDNA variants for their association with survival and QoL outcomes in critically ill patients. These results revealed distinct genomic signals linked to both mortality and poor functional recovery. Three variants reaching statistical significance discriminated survivors from non-survivors; rs3928305 (a synonymous *MT-SHLP3* variant) notably demonstrating a strong association with survival (10:1 ratio, *p* = 0.033). *MT-SHLP3* has been reported to have cytoprotective and anti-apoptotic function [50, 51], raising the possibility that variants in this gene could influence resilience or recovery in critically ill patients.

Five variants were significantly associated with overall outcomes defined as death or survival with poor physical function. Two variants in the control region have been linked to reduced VO2 max [40, 41], suggesting potential relevance to mitochondrial fitness and systemic resilience. Additionally, protein-coding *MT-ND5* variants, including a synonymous change m.12,612A*>*G previously reported to associate with coronary artery disease [42] and a missense variant m.13,708G*>*A previously implicated in kidney dysfunction [43] and neurodegenerative disorders [39], were enriched among poor-outcome patients. These findings highlight the possible influence of mitochondrial genetic variation on clinical trajectories in critical illness and support further investigation into their mechanistic and prognostic roles.

Despite the advances described in this report, several limitations remain. First, while restricting analysis to 75–149 bp fragments minimizes NUMT contamination and improves mtDNA specificity, it excludes longer inserts that may carry biologically relevant signals, potentially reducing sensitivity to certain DAMP species. Second, inserts in the 150–224 bp range, which likely include mononucleosome-associated NUMTs, present interpretive challenges. While in our study, in most of our patients, the range displays a high mtDNA-to-NUMT ratio. Nevertheless, residual NUMT contamination cannot be ruled out, and NUMT burden may vary by patient or condition being studied thus limiting generalizability of the size restriction approach. Third, the non-Gaussian distribution of mtDNA DAMPs observed in this ARF patient population, which skews toward non-survivors, complicates the use of abundance as a predictive biomarker. This skew may reflect biological heterogeneity or limited sample size and challenges the application of uniform clinical thresholds. Fourth, and related to variant detection, while the mismatch percentage effectively filters NUMT artifacts, its reliability depends on accurate reference alignments and may still misclassify low-frequency heteroplasmies, especially those from polymorphic NUMTs absent in the reference genome. Finally, the small sample size and lack of serial tissue sampling limit conclusions about the clinical or mechanistic significance of variants associated with ARF outcomes. However, our findings support the utility of our approach for identifying mitochondrial variants.

In closing, this study describes an improved, comprehensive approach to the analysis of circulating mtDNA DAMPs in critically ill patients by addressing the confounding issue of NUMT contamination. Through insert-size stratification and the development of the read mismatch percentage metric, we have enhanced the accuracy of mtDNA quantification and variant detection in liquid biopsies. Conflicting results in prior studies regarding mtDNA DAMP parameters [13] have been ascribed to differences in methods, patient selection, and sample handling. Our approach mitigates many of these issues by employing cost-effective, reproducible methods that offer standardized insight into mtDNA-derived signals [18]. Our findings reinforce the prognostic relevance of mtDNA abundance and heteroplasmic signatures in acute and post-acute outcomes, suggesting that mitochondrial dysfunction plays a dynamic role across the critical illness trajectory. While limitations persist, particularly regarding residual NUMT interference and small cohort size, this work lays the foundation for future studies aimed at refining mitochondrial biomarkers and integrating them into precision medicine strategies for critical care.

## Conclusions

Standard qPCR-based approaches to study mtDNA DAMPs suffer from numerous limitations. Firstly, they rely on pre-selected amplicon designs which are not standardized in the field. This invites uncertainty, as the putative innate immune receptors acted on by mtDNA have known sequence specificity. Polymorphic and somatic NUMT continue to be reported and are especially problematic for qPCR amplicon design, as they are not contained in the reference genome. Utilizing the high mtDNA coverage provided by targeted mtDNA sequencing technique, we identified polymorphic NUMT in ARF patient samples, allowing us to mitigate their technical impact *in silico*. NUMT-mitigated abundance and DNA insert size showed stronger associations with non-survival than their non NUMT-mitigated counterparts. Additionally, variants associated with ARF outcome, opening the possibility that this method could provide a similar patient-specific liquid biopsy to those utilized in the cancer field. This workflow, which utilizes only commercially-available reagents and open source software, supports further studies on serum mtDNA variants to predict acute and long-term outcomes in critical illness.

## Methods

### DNA Isolation, Enrichment, and ILLUMINA Sequencing

DNA was isolated from serum, Illumina libraries prepared, and mitochondrial DNA captured with MyBaits RNA bait capture kit as described in [18].150 bp paired end sequencing was performed by Novogene on an Illumina HiSeq 4000 for the acute study and Hi-Seq X for the QoL study.

### Bioinformatics Processing

Data were processed through the bioinformatics pipeline introduced in Daly et al., 2022 [18]. In short, raw paired end fastq files were adapter trimmed with Cutadapt, and then mapped to the GRCh38 human reference genome with the BWA MEM. Custom post-processing programs called the coverage, defined as the number of sequenced and aligned bases meeting mapping quality 20 which cover a reference base. Mitochondrial coverage was normalized to nuclear DNA (nDNA), defined as mean NUMT coverage. Fragment size cannot be directly measured by standard paired-end Illumina sequencing due to blunting of 3’ and 5’ overhangs during library preparation end repair and the fact that mate pairs may be aligned chimerically, so, we used the insert size field from paired-end “bam” files to generate curves for reads aligned to annotated NUMTs and the mitochondrial genome [52, 53]. Insert was size assessed as a histogram plotting the percentage of observed fragment sizes from zero to 1,000, with the assumption that Illumina sequencing instruments don’t efficiently cluster inserts past 1,000 bp, and reads required a minimum mapping quality 20. A 5 bp median smoothing function was employed with the assumption that these spikes may be some type of alignment or batch artifact (see results for details). Finally, base pileups of the mitochondria generated with BCFtools mpileup with maximum depth 10k, minimum base quality 20, and minimum mapping quality 20; and further processed to call variants with a custom BCFtools plugin called “heteroplasmy”, hosted in a forked GitHub repository for BCFtools. To be considered a homoplasmic variant, alleles required 10 reads on the forward and reverse strand and 99 % VAF. In addition to the 10 reads mapping to both the forward and reverse strand, a minimum depth of 500 and minimum VAF 0.01 were required to call a heteroplasmic variant. Full details of the pipeline are available in Daly et al., 2022 [18]; the supplementary methods section of which details the source code repository https://github.com/GrantDaly/mitochondrial-alignments. Additional filters were added to address highly homologous polymorphic NUMT, as described in Results. Code is available in the same Github repository as “identifynumt.cpp”. The software iterates through all reads aligned to the mitochondria and calculates the percentage of A,T,C, or G bases from reads where the mate pair read was aligned to another contig.

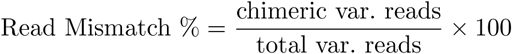

### Modeling insert size parameters

Mitochondrial and NUMT insert size distributions were recorded as arrays ranged 1-1,000 bp, recording the number of inserts observed at each size in the range. A 5 bp median smoothing function was used for plotting the histograms, which visually reduced 1 bp spikes we reasoned to be technical artifacts. This, however, was not sufficient to remove apparent NUMT contaminants, which displayed similar widths to reads mapped to known NUMT. As the mitochondrial inserts appeared to have broad peaks with a long right tail, the Skew Student’s t distribution was adapted, and data were fit with Scipy v1.14.0 Differential Evolution algorithm with parameters described in Table S2 [54, 55]. NUMT were similarly modeled with Differential Evolution, but using the Generalized Normal distribution from Scipy, and fitting for both putative mononucleosomes (abbreviated as “mono” or “mononuc”) and dinucleosomes (abbreviated as “dinuc”). The mode modeled insert size was chosen as the point statistic for group comparisons, with the rationale it was a good point measure for the highly right-skewed mtDNA and also summarized the sharp mononucleosome peaks and less sharp but symmetrical dinucleosome peaks well. A Monte Carlo simulation technique was employed to estimate general parameters of the three peaks, wherein 10,000 samples with replacement were taken of the mode modeled inserts, and the 2.5 %, 50 %, and 97.5 % quantiles were taken to generate a confidence interval as well as a measure of central tendency of the insert distributions.

### Bayesian modeling

The Bayesian Estimation Supersedes the T-Test (BEST) strategy was employed to test mean differences of normalized mitochondrial coverage and abundance (mean mtDNA*/*mean NUMT) and raw and modeled mean insert size [29]. This strategy offers numerous advantages including robustness to outliers, greater ability to test assumptions such as normality and equivalence of variance, as well as to extend the model to other distributions. For each model, posterior distributions were estimated through the Python PyMC v5.16.2 default No-U-Turn Sampler with 10 chains of 1,000 tuning draws and 10,000 draws [56]. In this analysis, the modeled mean insert size was modeled with the Student’s t distribution, whereas the more skewed normalized coverage data were log transformed and fit with a normal distribution. The objective of the model fitting was to find the 95 % highest density interval (HDI) for the mean (or in the case of coverage the mean of the log transformed data) of each group. Once fitted, differences could be evaluated between the group means and the HDI of these differences could themselves be taken. A 95 % HDI different than 0 was the criterion to call differences between groups.

Table S4 displays the parameters used to model survival abundance. Note that all data was log transformed before being introduced to the model, so mean *µ* is really the mean of the natural logarithm of abundance. Also, a *µ* and *τ* and *σ* term was fit for each measure of abundance: coverage, all inserts abundance, 75-149, 150-224, 225-299, & 300-374 bp abundances as well as for survival and non-survival for a total of 6 *×* 2 = 12 *µ* and 12 *τ* and *σ* terms. Finally, a Normal likelihood function was chosen with parameter distributions *µ* and *τ*. Table S6 displays survival raw and fitted insert size, which was constructed in a similar manner, with the major difference that it used the PyMC StudentT likelihood distribution, which additionally included a “nu” normality parameter.

For the QoL data a subset of patients had admission and discharge samples available. We accounted for repeated measures by modeling samples from the same patient as coming from a multivariate distribution with a correlation term. As only 28/49 patients had both admission and discharge samples available (Table S1b), the Bayesian model fitting imputed missing values based on the observed correlation between available patients’ admission and discharge samples. The correlation term was designed to favor values closer to the available sample for the patient, and thus may underestimate differences between admission and discharge. As can be seen in Table S5 and S7, similar BEST models were constructed with the multivariate Normal likelihood function for abundance and the multivariate Student’s t likelihood function for QoL, each of which were parameterized with PyMC’s Cholesky distribution to model the correlation between admission and discharge samples.

## List of abbreviations

ARF: acute respiratory failure
mtDNA: mitochondrial DNA
nDNA: nuclear DNA
DAMP: damage-associated molecular pattern
bp: base pair
TLR9: Toll-like receptor 9
qPCR: Quantitative real-time PCR
mononuc: mononucleosome
dinuc: dinucleosome
BEST: Bayesian Estimation Supersedes the T-Test

## Declarations

### Ethics approval and consent to participate

This was a secondary analysis of samples collected for study NCT00976833, a randomized clinical trial evaluating early physical therapy in patients with acute respiratory failure (ARF). The Institutional Review Board of the enrolling hospital, Wake Forest Baptist Medical Center, North Carolina, gave ethical approval for patient sample collection and analysis.

### Consent for publication

Not applicable.

### Availability of data and materials

All data and analysis code produced for the present study, including raw sequencing reads, will be made publicly available upon final manuscript publication. Pending publication, details or data may be shared upon reasonable requests.

Workflow Description Language (WSL) pipelines were hosted on Dockstore (https://dockstore.org/search?organization=GrantDaly&entryType=workflows&search=grant%20daly).

Any additional information required to reanalyze the data reported in this paper is available from the lead contact upon request.

### Authors’ contributions

G.T.D. developed bioinformatics software and statistical models, and generated the figures. V.M.P. performed protocols for patient serum DNA isolation, HTS library preparation, and mtDNA enrichment protocol. M.S.M. and A.I.H. contributed to statistical analyses and reporting of results. J.T.R. contributed to the bioinformatics analysis design. D.C.F. and P.E.M. designed, conducted, and secured funding for the clinical trial, and analyzed clinical outcomes. L.D.P. was the clinical research coordinator for the original study, and provided samples, clinical data, and deidentified data for the present study. G.T.D., E.M.H., M.N.G., and R.J.L. drafted the manuscript. M.N.G. and R.J.L. conceived the design, secured funding, and analyzed results from the study. All authors reviewed the manuscript.

## References

[1] Langley, R. J. & Wong, H. R. Early diagnosis of sepsis: is an integrated omics approach the way forward? Mol. Diagn. Ther. 21, 525–537 (2017).

[2] Langley, R. J. et al. An integrated clinico-metabolomic model improves prediction of death in sepsis. Sci. Transl. Med. 5, 195ra95 (2013).

[3] Langley, R. J. et al. Integrative “Omic” Analysis of Experimental Bacteremia Identifies a Metabolic Signature That Distinguishes Human Sepsis from Systemic Inflammatory Response Syndromes. Am. J. Respir. Crit. Care Med. 190, 445–455 (2014).

[4] Singer, M. Mitochondrial function in sepsis: Acute phase versus multiple organ failure. Crit. Care Med. 35, S441 (2007).

[5] Singer, M. The role of mitochondrial dysfunction in sepsis-induced multi-organ failure. Virulence 5, 66–72 (2014).

[6] Dobson, A. W. et al. Enhanced mtDNA repair capacity protects pulmonary artery endothelial cells from oxidant-mediated death. Am. J. Physiol. Lung Cell Mol. Physiol. 283, L205–L210 (2002).

[7] Grishko, V., Solomon, M., Wilson, G. L., LeDoux, S. P. & Gillespie, M. N. Oxygen radical-induced mitochondrial DNA damage and repair in pulmonary vascular endothelial cell phenotypes. Am. J. Physiol. Lung Cell Mol. Physiol. 280, L1300–L1308 (2001).

[8] Ruchko, M. et al. Mitochondrial DNA damage triggers mitochondrial dysfunction and apoptosis in oxidant-challenged lung endothelial cells. Am. J. Physiol. Lung Cell Mol. Physiol. 288, L530–L535 (2005).

[9] Tao, G. et al. Advances in crosstalk among innate immune pathways activated by mitochondrial DNA. Heliyon 10, e24029 (2024).

[10] Zhang, Q. et al. Circulating mitochondrial DAMPs cause inflammatory responses to injury. Nature 464, 104–107 (2010).

[11] Zhang, Q., Itagaki, K. & Hauser, C. J. Mitochondrial DNA is released by shock and activates neutrophils via p38 map kinase. Shock 34, 55–59 (2010).

[12] Kuck, J. L. et al. Mitochondrial DNA damage-associated molecular patterns mediate a feed-forward cycle of bacteria-induced vascular injury in perfused rat lungs. Am. J. Physiol. Lung Cell Mol. Physiol. 308, L1078–1085 (2015).

[13] Harrington, J. S. et al. Circulating mitochondrial DNA as predictor of mortality in critically ill patients: a systematic review of clinical studies. Chest 156, 1120–1136 (2019).

[14] Wei, W. et al. Nuclear-embedded mitochondrial DNA sequences in 66,083 human genomes. Nature 611, 105–114 (2022).

[15] Zhou, W. et al. Somatic nuclear mitochondrial DNA insertions are prevalent in the human brain and accumulate over time in fibroblasts. PLoS Biol. 22, e3002723 (2024).

[16] Bruhm, D. C. et al. Genomic and fragmentomic landscapes of cell-free DNA for early cancer detection. Nat. Rev. Cancer 25, 341–358 (2025).

[17] Ries, M. et al. Identification of novel oligonucleotides from mitochondrial DNA that spontaneously induce plasmacytoid dendritic cell activation. J. Leukoc. Biol. 94, 123–135 (2013).

[18] Daly, G. T. et al. Novel attributes of cell-free plasma mitochondrial DNA in traumatic injury. Clin. Transl. Med. 12, e1055 (2022).

[19] van der Pol, Y. et al. The landscape of cell-free mitochondrial DNA in liquid biopsy for cancer detection. Genome Biol. 24, 229 (2023).

[20] Wake Forest University. Standardized rehabilitation for ICU patients with acute respiratory failure. Clinical trial registration NCT00976833, clinicaltrials.gov (2018).

[21] Gandotra, S. et al. Physical function trajectories in survivors of acute respiratory failure. Annals ATS 16, 471–477 (2019).

[22] Morris, P. E. et al. Standardized rehabilitation and hospital length of stay among patients with acute respiratory failure: a randomized clinical trial. JAMA 315, 2694–2702 (2016).

[23] Jiang, P., et al. Lengthening and shortening of plasma DNA in hepatocellular carcinoma patients. Proc. Natl. Acad. Sci. U.S.A. 112, E1317–E1325 (2015).

[24] Lo, Y. M. D. et al. Maternal plasma DNA sequencing reveals the genome-wide genetic and mutational profile of the fetus. Sci. Transl. Med. 2, 61ra91 (2010).

[25] Zhang, R., Nakahira, K., Guo, X., Choi, A. M. & Gu, Z. Very short mitochondrial DNA fragments and heteroplasmy in human plasma. Sci. Rep. 6, 36097 (2016).

[26] Shi, J., Zhang, R., Li, J. & Zhang, R. Size profile of cell-free DNA: a beacon guiding the practice and innovation of clinical testing. Theranostics 10, 4737–4748 (2020).

[27] Harrison, R. L. Introduction To Monte Carlo Simulation. AIP Conf. Proc. 1204, 17–21 (2010).

[28] Piantadosi, C. A. Mitochondrial DNA, oxidants, and innate immunity. Free Radic. Biol. Med. 152, 455–461 (2020).

[29] Kruschke, J. K. Bayesian Estimation Supersedes the T test. J. Exp. Psychol. Gen. 142, 573–603 (2013).

[30] Inoue, S. et al. Post-intensive care syndrome: its pathophysiology, prevention, and future directions. Acute Med. Surg. 6, 233–246 (2019).

[31] Galvis-Pedraza, M., Beumeler, L. F. E., van der Slikke, E. C., Boerma, E. C. & van Zutphen, T. Mitochondrial DNA in plasma and long-term physical recovery of critically ill patients: an observational study. Intensive Care Med. Exp. 12, 99 (2024).

[32] Hawkins, R. B. et al. Persistently increased cell-free DNA concentrations only modestly contribute to outcome and host response in sepsis survivors with chronic critical illness. Surgery 167, 646–652 (2020).

[33] Corsonello, A. et al. Prognostic significance of the Short Physical Performance Battery in older patients discharged from acute care hospitals. Rejuvenation Res. 15, 41–48 (2012).

[34] de Fátima Ribeiro Silva, C., Ohara, D. G., Matos, A. P., Pinto, A. C. P. N. & Pegorari, M. S. Short Physical Performance Battery as a measure of physical performance and mortality predictor in older adults: a comprehensive literature review. Int. J. Environ. Res. Public Health 18, 10612 (2021).

[35] Singh, K. K., Choudhury, A. R. & Tiwari, H. K. Numtogenesis as a mechanism for development of cancer. Semin. Cancer. Biol. 47, 101–109 (2017).

[36] Dayama, G., Emery, S. B., Kidd, J. M. & Mills, R. E. The genomic landscape of polymorphic human nuclear mitochondrial insertions. Nucleic Acids Res. 42, 12640–12649 (2014).

[37] Thorvaldsdóttir, H., Robinson, J. T. & Mesirov, J. P. Integrative Genomics Viewer (IGV): high-performance genomics data visualization and exploration. Brief. Bioinform. 14, 178–192 (2013).

[38] McLaren, W. et al. The Ensembl Variant Effect Predictor. Genome Biol. 17, 122 (2016).

[39] Lott, M. T. et al. mtDNA variation and analysis using Mitomap and Mitomaster. Curr. Protoc. Bioinformatics 44, 1.23.1–26 (2013).

[40] Martínez-Redondo, D., et al. Human mitochondrial haplogroup H: The highest VO2max consumer – is it a paradox? Mitochondrion 10, 102–107 (2010).

[41] Vellers, H. L. et al. Association between mitochondrial DNA sequence variants and V̇O_2_ max trainability. Med. Sci. Sports Exerc. 52, 2303–2309 (2020).

[42] Vilne, B., Sawant, A. & Rudaka, I. Examining the association between mitochondrial genome variation and coronary artery disease. Genes (Basel) 13, 516 (2022).

[43] Cañadas-Garre, M., et al. Genetic variants affecting mitochondrial function provide further insights for kidney disease. BMC Genomics 25, 576 (2024).

[44] Cloonan, S. M. & Choi, A. M. Mitochondria in lung disease. J. Clin. Invest. 126, 809–820 (2016).

[45] Duran-Bedolla, J. et al. Sepsis, mitochondrial failure and multiple organ dysfunction. Clin. Investig. Med. 37, E58–E69 (2014).

[46] Kellner, M. et al. in ROS signaling in the pathogenesis of Acute Lung Injury (ALI) and Acute Respiratory Distress Syndrome (ARDS) (ed. Wang, Y. X.) Pulmonary Vasculature Redox Signaling in Health and Disease 105–137 (Springer International Publishing, Cham, Switzerland, 2017).

[47] Xue, L., Moreira, J. D., Smith, K. K. & Fetterman, J. L. The mighty NUMT: mitochondrial DNA flexing its code in the nuclear genome. Biomolecules 13, 753 (2023).

[48] Liu, Y. et al. NGS-based accurate and efficient detection of circulating cell-free mitochondrial DNA in cancer patients. Mol. Ther. Nucleic Acids 23, 657–666 (2021).

[49] Lou, C. et al. The mtDNA fragments within exosomes might be novel diagnostic biomarkers of non-small cell lung cancer. Pathol. Res. Pract. 249, 154718 (2023).

[50] Cobb, L. J. et al. Naturally occurring mitochondrial-derived peptides are age-dependent regulators of apoptosis, insulin sensitivity, and inflammatory markers. Aging 8, 796–809 (2016).

[51] Yen, K., Miller, B., Kumagai, H., Silverstein, A. & Cohen, P. Mitochondrial-derived microproteins: from discovery to function. Trends Genet. 41, 132–145 (2025).

[52] Li, H. et al. The Sequence Alignment/Map format and SAMtools. Bioinformatics 25, 2078–2079 (2009).

[53] Turner, F. S. Assessment of insert sizes and adapter content in fastq data from NexteraXT libraries. Front. Genet. 5, 5 (2014).

[54] Jones, M. C. & Faddy, M. J. A skew extension of the t-distribution, with applications. J. R. Stat. Soc. Series B Stat. Methodol. 65, 159–174 (2003).

[55] Virtanen, P. et al. SciPy 1.0: fundamental algorithms for scientific computing in Python. Nat. Methods 17, 352 (2020).

[56] Vincent, B. T., et al. pymc-devs/pymc-examples: December 2022 snapshot. Zenodo (2022).

