## Supplemental Tables and Figures for "Serum mtDNA DAMP abundance, fragmentation and heteroplasmic variants associate with Acute Respiratory Failure outcome: A secondary analysis of study NCT00976833"

**Table S1.** Samples sequenced for survival and quality of life studies.

A) Survival

| | Survival | Nonsurvival | All |
|---|---|---|---|
| # of Patients | 36 | 36 | 72 |

B) QoL

| Samples Available QoL | Adm. & Dis. | Adm., No Dis. | Dis., No Adm. | Total |
|---|---|---|---|---|
| Good | 12 | 6 | 5 | 23 |
| Poor | 16 | 2 | 8 | 26 |
| Total | 28 | 8 | 13 | 49 |

**Table S2.** Distributions used to fit Mitochondrial and NUMT insert size.

A) Skew T distribution pdf for mitochondria

$$pdf_{skew\_t}(t,a,b) = C\_skew\_t_{a,b}^{-1}\{1 + t/(a+b+t^2)^{1/2}\}\{1 - t/(a+b+t^2)^{1/2}\}$$
$$C\_skew\_t = 2^{a+b-1}B(a,b)(a+b)^{\frac{1}{2}}$$
$$pdf_{skew\_t\_loc}(t,a,b,loc,scale) = pdf_{skew\_t}((t-loc)/scale,a,b)/scale$$

B) Skew T parameter ranges for differential evolution fit

$$pdf_{skew\_t\_loc}(t;a,b,loc,scale) = t \in [sampleDependent], a \in [1,20], b \in [1,20], loc \in [50,150], scale \in [1/100,100]$$

C) Generalized normal distribution pdf for NUMT

$$pdf_{gen\_norm}(x,b) = \frac{\beta}{2\Gamma(1/\beta)}exp(-|x|^{\beta})$$
$$pdf_{gen\_norm\_loc}(x,b,loc,scale) = pdf_{gen\_norm}((x-loc)/scale,b)/scale$$

D) Generalized normal parameter ranges for differential evolution fit

$$pdf_{gen\_norm\_loc\_mono}(x,b,loc,scale) = x \in [100,250], b \in [1,2], loc \in [30,70], scale \in [1/10,100]$$
$$pdf_{gen\_norm\_loc\_dinuc}(x,b,loc,scale) = x \in [250,450], b \in [1,2], loc \in [30,70], scale \in [1/10,100]$$

**Table S3.** Bootstrap estimation of insert size quantiles 10%,50%, and 90% for mitochondria, NUMT mononucleosome, and NUMT dinucleosome. Data reported are the median value of all 10K draws.

| | 10% | 50% | 90% |
|---|---|---|---|
| Mito. | 115 | 187 | 325 |
| Mono. | 145 | 167 | 189 |
| Dinuc. | 272 | 319 | 366 |

**Table S4.** Model Specification for Survival abundance

$$\begin{aligned}
\text{precision multiplier} &= 0.1\\
\text{sigma multiplier} &= 10\\
\text{sample mean} &= \text{Mean}(\log(\text{Data}))\\
\text{sample SD} &= \text{Stdev}(\log(\text{Data}))\\
\text{sample tau} &= 1/\text{sample SD}^2\\
\sigma &\sim \text{Uniform}(\text{sample SD}/\text{sigma multiplier}), \text{sample SD} \times \text{sigma multiplier})\\
\tau &\sim \text{Deterministic}(1/\sigma)\\
\mu_{\text{Survival,Non-survival}} &\sim \text{Normal}(\text{mu} = \text{sample mean}, \text{tau} = \text{precision multiplier} \times \text{sample tau})\\
\log(\text{abundance}) &\sim \text{Normal}(mu = \mu[\text{Condition}], tau = \tau[\text{Condition}])
\end{aligned}$$

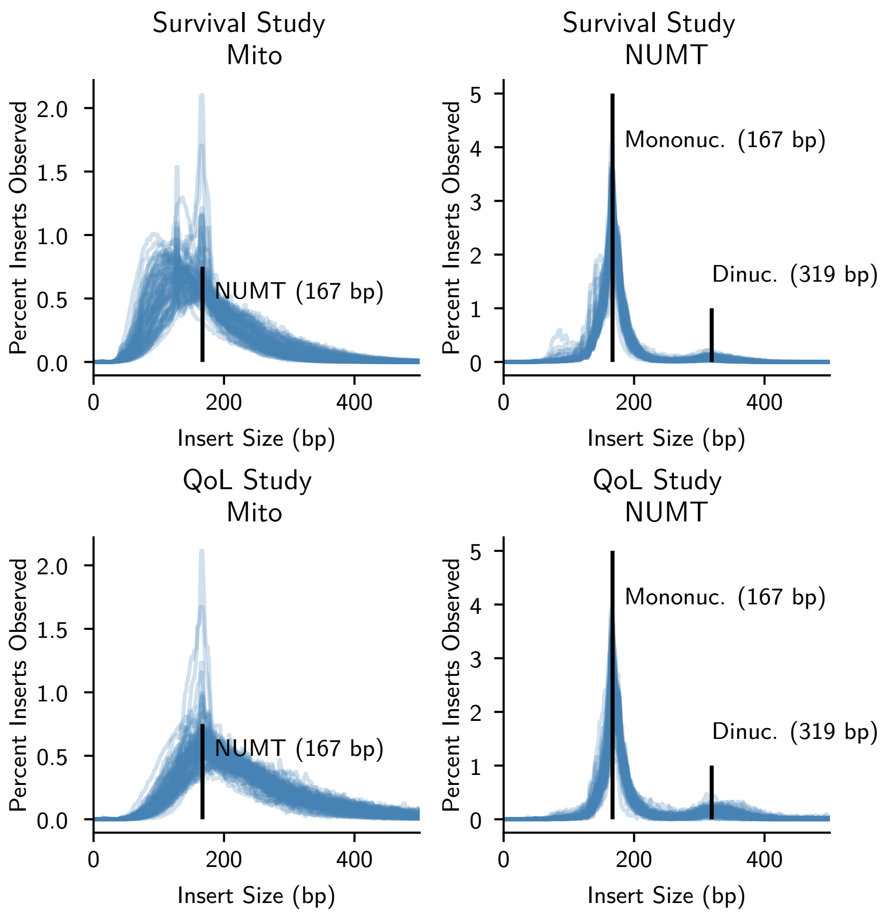


**Figure S1.** Median smoothed insert size histograms

**Table S5.** Model Specification for QoL abundance

$$
\begin{aligned}
\text{precision multiplier} &= 0.1 \\
\text{sigma multiplier} &= 10 \\
\text{sample mean} &= \mathrm{Mean}(\log(\text{Data})) \\
\text{sample SD} &= \mathrm{Stdev}(\log(\text{Data})) \\
\text{sample tau} &= 1/\text{sample SD}^2 \\
\sigma &\sim \mathrm{Uniform}(\text{sample SD}/\text{sigma multiplier}, \text{sample SD} \times \text{sigma multiplier}) \\
\tau &\sim \mathrm{Deterministic}(1/\sigma) \\
\text{Cholesky} &\sim \mathrm{LKJCholeskyCov}(\text{eta} = 2, \text{n} = 2(\text{For Adm./Dis.}), \text{sd\_dist} = \mathrm{Exponential}(0.5)) \\
\mu_{\text{QoL} \times \text{Time}} &\sim \mathrm{Normal}(\text{mu} = \text{sample mean}, \text{tau} = \text{precision multiplier} \times \text{sample tau}) \\
\log(\text{abundance}) &\sim \mathrm{MVNormal}(\text{mu} = \mu[\text{QoL, Time}], \text{chol} = \text{Cholesky})
\end{aligned}
$$

**Table S6.** Model Specification for Survival raw and fitted insert size.

$$
\begin{aligned}
\text{precision multiplier} &= 0.000001 \\
\text{sigma multiplier} &= 1,000 \\
\text{sample mean} &= \mathrm{Mean}(\log(\text{Data})) \\
\text{sample SD} &= \mathrm{Stdev}(\log(\text{Data})) \\
\text{sample tau} &= 1/\text{sample SD}^2 \\
\nu &= \mathrm{Exponential}(\text{lam} = 1/29) + 1 \\
\sigma &\sim \mathrm{Uniform}(\text{sample SD}/\text{sigma multiplier}, \text{sample SD} \times \text{sigma multiplier}) \\
\tau &\sim \mathrm{Deterministic}(1/\sigma) \\
\mu_{\text{Survival,Non-survival}} &\sim \mathrm{Normal}(\text{mu} = \text{sample mean}, \text{tau} = \text{precision multiplier} \times \text{sample tau}) \\
\text{insert size} &\sim \mathrm{StudentT}(\text{mu} = \mu[\text{Condition}], \text{sigma} = \sigma[\text{Condition}], \text{nu} = \nu)
\end{aligned}
$$

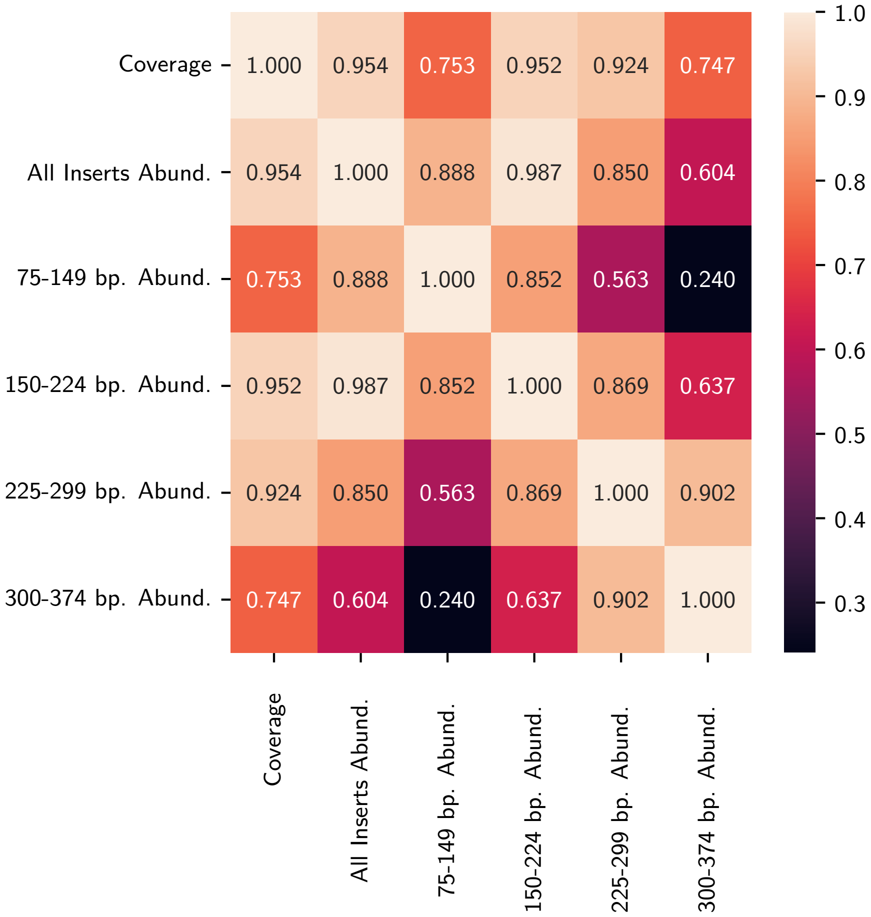


**Figure S2.** Spearman correlation $\rho$ between mitochondrial coverage and insert size abundances for the denoted size ranges, all normalized to NUMT.

**Table S7.** Model Specification for QoL raw and fitted mean insert size.

$$
\begin{aligned}
\text{precision multiplier} &= 0.1 \\
\text{sigma multiplier} &= 10 \\
\text{sample mean} &= \mathrm{Mean}(\log(\text{Data})) \\
\text{sample SD} &= \mathrm{Stdev}(\log(\text{Data})) \\
\text{sample tau} &= 1/\text{sample SD}^2 \\
\sigma &\sim \mathrm{Uniform}(\text{sample SD}/\text{sigma multiplier}), \text{sample SD} \times \text{sigma multiplier}) \\
\tau &\sim \mathrm{Deterministic}(1/\sigma) \\
\text{Cholesky} &\sim \mathrm{LKJCholeskyCov}(\text{eta} = 2, \text{n} = 2(\text{For Adm./Dis.}), \text{sd\_dist} = \mathrm{Exponential}(0.5)) \\
\mu_{\text{QoL}\times\text{Time}} &\sim \mathrm{Normal}(\text{mu} = \text{sample mean}, \text{tau} = \text{precision multiplier} \times \text{sample tau}) \\
\text{insert size} &\sim \mathrm{MVStudentT}(\text{mu} = \mu[\text{QoL, Time}], \text{nu} = \nu, \text{chol} = \text{Cholesky})
\end{aligned}
$$

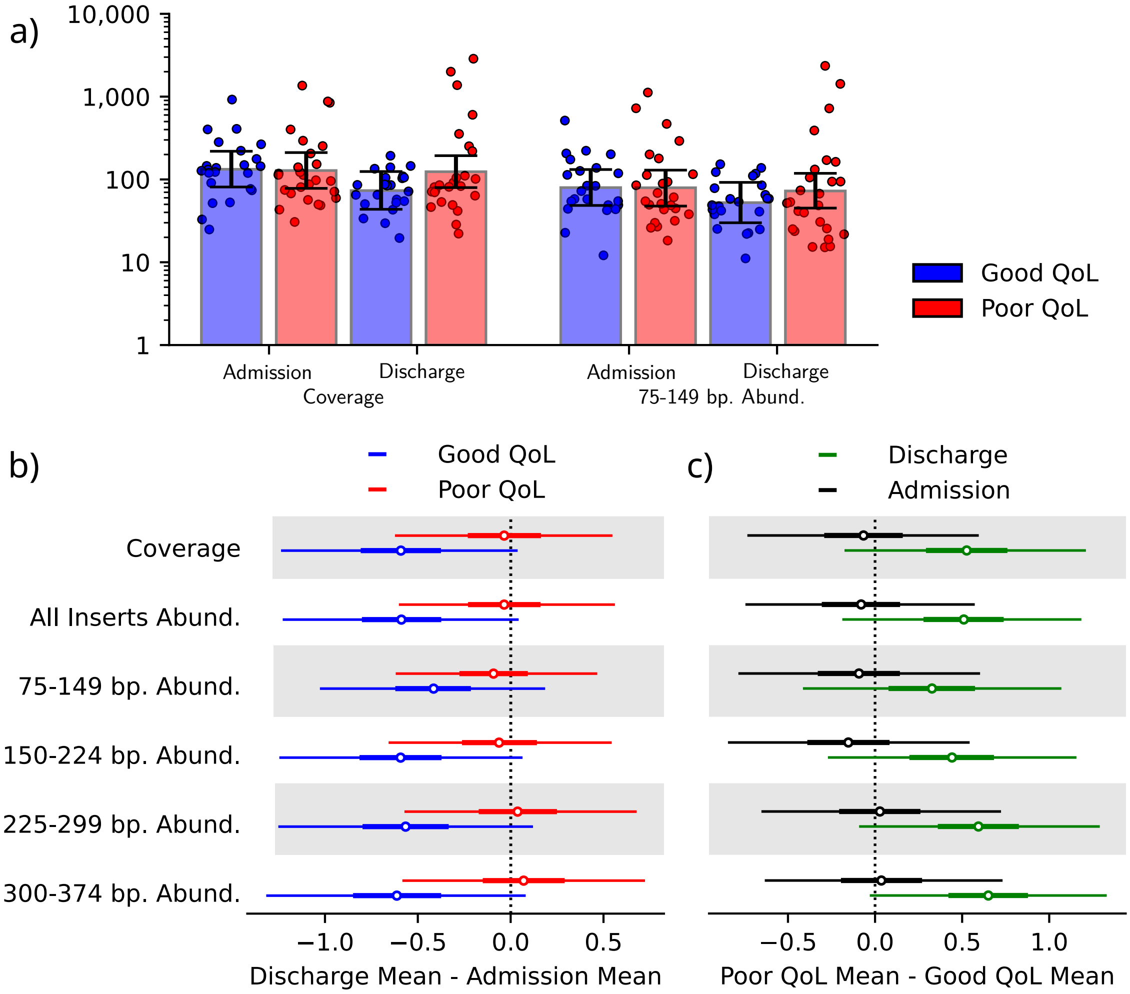


**Figure S3.** mtDNA abundance and coverage by QoL and time. (A) Points and 95% HDI displayed for coverage and 75-149 bp. inserts. 95% HDI of differences in abundance of denoted size range or coverage by (B) time (discharge - admission) or (C) QoL (poor - good). Difference "0" is displayed as dotted vertical line.

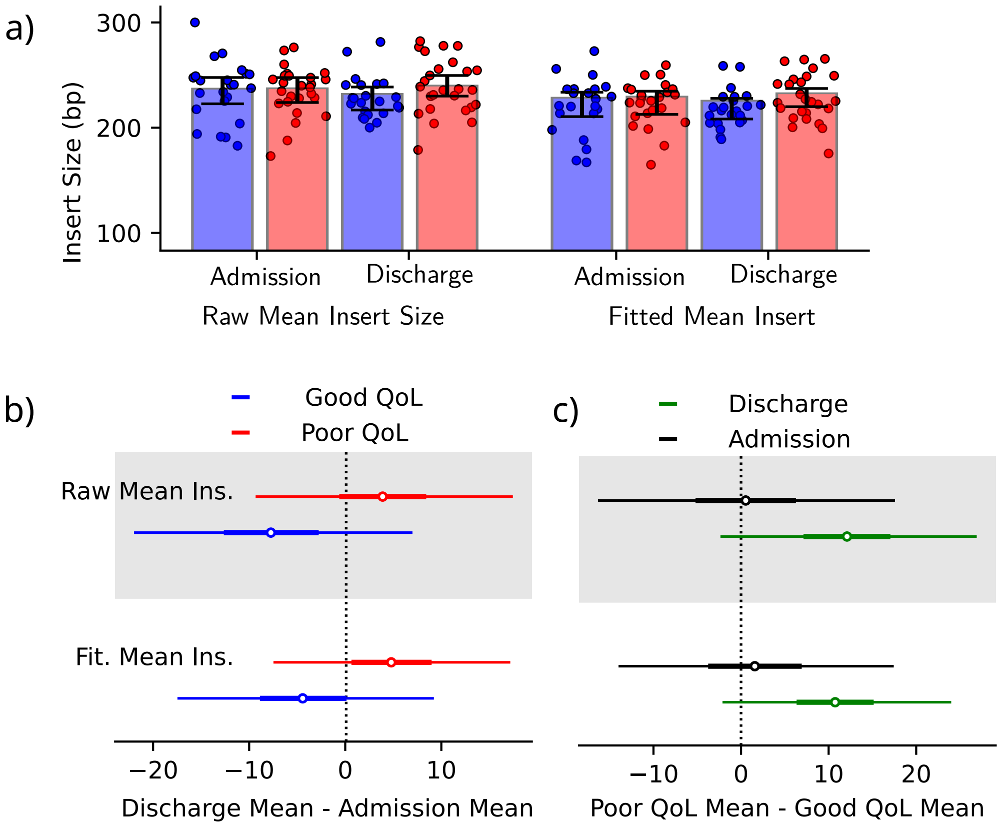


**Figure S4.** Raw and fitted mean insert size by QoL and time. A) Points and 95% HDI for raw and fitted mean insert size. 95% HDI of insert size differences by (B) time (discharge - admission), and (C) QoL (poor QoL - good QoL). Difference "0" displayed as dotted vertical line.

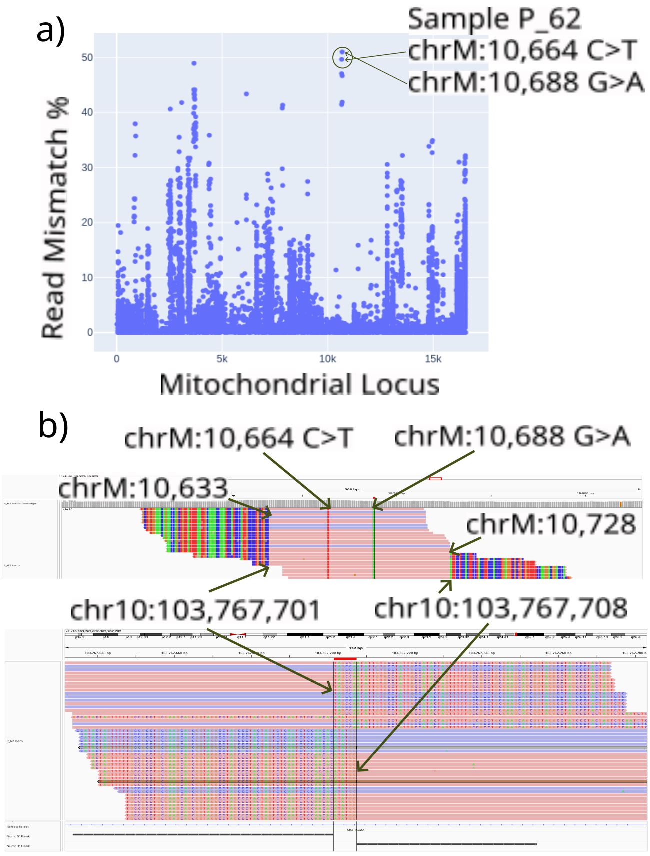


**Figure S5.** Mismatch percentage for read pairs mapping to the mitochondrial and nuclear genomes. (A) Patient variant allele mismatch percentages by mitochondrial position. (B) Representative patient sample polymorphic NUMT.

**Table S8.** Quantiles for VAF and read mismatch %.

| Quantile | VAF | Read Mismatch % |
|---:|---:|---:|
| 0.1 | 1.6 | 0.0 |
| 0.2 | 2.6 | 0.2 |
| 0.3 | 4.2 | 0.4 |
| 0.4 | 7.5 | 0.8 |
| 0.5 | 13.2 | 1.3 |
| 0.6 | 25.3 | 2.0 |
| 0.7 | 86.1 | 3.1 |
| 0.8 | 97.3 | 4.9 |
| 0.9 | 99.4 | 9.3 |

**Table S9.** Variant associations by permutation test comparing the variants observed at high VAf in survival non-survival or survival with poor QoL and non-survival compared to survival with good QoL.

A) Survival

| HGVSG | dbSNP | Ratio | p-value | Annotations |
|---|---|---:|---:|---:|
| NC_012920.1:m.1719G>A | rs3928305 | 10:1 | 0.023 | MT-SHLP3 Syn. |
| NC_012920.1:m.16187C>T | rs879041967 | 5:0 | 0.032 | MT-CR, lung tumor |
| NC_012920.1:m.10589G>A | rs2853487 | 5:0 | 0.032 | MT-ND4L Syn. |

B) Non-Survival or Poor QoL

| HGVSG | dbSNP | Ratio | p-value | Annotations |
|---|---|---:|---:|---:|
| NC_012920.1:m.295C>T | rs41528348 | 7:0 | 0.020 | MT-CR, Low $VO_2$ max |
| NC_012920.1:m.12612A>G | rs28359172 | 6:0 | 0.024 | MT-ND5 Syn. |
| NC_012920.1:m.462C>T | rs41402146 | 5:0 | 0.032 | MT-CR, Low $VO_2$ max |
| NC_012920.1:m.16172T>C | rs2853817 | 5:0 | 0.044 | MT-CR |
| NC_012920.1:m.13708G>A | rs28359178 | 7:1 | 0.047 | MT-ND5 Mis., PD-ADS |
